# Diagnostic Efficacy of Rapid Antigen Testing for SARS-CoV-2: The COVid-19 AntiGen (COVAG) study

**DOI:** 10.1101/2021.08.04.21261609

**Authors:** Christoph Wertenauer, Geovana Brenner Michael, Alexander Dressel, Caroline Pfeifer, Ulrike Hauser, Eberhard Wieland, Christian Mayer, Caren Mutschmann, Martin Roskos, Hans-Jörg Wertenauer, Winfried März

## Abstract

**Background:** Widely available rapid testing is pivotal to the fight against COVID-19. Real-time reverse transcription-polymerase chain reaction (rRT-PCR) remains the gold standard. We compared two frequently used commercial rapid diagnostic tests (RDTs) for SARS-CoV-2-antigens, the SD Biosensor SARS-CoV-2 Rapid Antigen Test (Roche Diagnostics) and the Panbio COVID-19 Ag Rapid Test (Abbott Diagnostics), against rRT-PCR for SARS-CoV-2 detection.

**Methods:** We compared the tests in 2215 all-comers at a diagnostic centre between February 1 and March 31, 2021. rRT-PCR-positive samples were examined for SARS-CoV-2 variants.

**Findings:** 338 participants (15%) were rRT-PCR-positive for SARS-CoV-2. The sensitivities of Roche-RDT and Abbott-RDT were 60.4% and 56.8% (P<0·0001) and specificities 99.7% and 99.8% (P=0·076), respectively. Sensitivity inversely correlated with rRT-PCR-derived Ct values. Unadjusted, the RDTs had higher sensitivities in individuals referred by treating physicians and health departments than those tested for other reasons, in persons without comorbidities compared to those with comorbidities, in individuals with symptoms suggesting COVID-19, and in the absence of SARS-CoV-2 variants compared to Alpha variant carriers. The associations of sensitivity with clinical symptoms and the SARS-CoV-2 genotype were robust against adjustment for Ct values. Assuming that 10 000 symptomatic individuals are tested, 500 of which are truly positive, the RDTs would generate 38 false-positive and 124 false-negative results. Assuming that 10 000 asymptomatic individuals are tested, including 50 true positives, 18 false-positives and 34 false-negatives would be generated.

**Interpretation:** The sensitivities of the two RDTs are unsatisfactory. This calls into question whether their widespread use is effective in the ongoing SARS-CoV-2 pandemic.

**Funding:** SYNLAB Holding Deutschland GmbH

**Research in context:** *Evidence before this study:* Small studies and a meta-analysis from the Cochrance collaboration indicate vastly different diagnostic efficacies of commercial rapid diagnostic tests (RDTs) for SARS-CoV-2 antigen. The impact of SARS-CoV-2 variants has not been known.

*Added value of this study:* This is one of the largest real-world studies of the diagnostic efficacy of two widely recommended RDTs SARS-CoV-2 antigen in comparison to rRT-PCR. The sensitivities of the two RDTs are unsatisfactory, mainly in asymptomatic persons. Presence of the SARS-CoV-2 Alpha Variant decreased both tests’ sensitivities significantly.

*Implications of all the available evidence:* Policy and health care providers should account for substantial limitations of RDTs for SARS-CoV-2 particular in asymptomatic persons. Research into alternative approaches to the screening for SARS-CoV-2 should be intensified.

## Introduction

On December 31, 2019, the Municipal Health Commission of Wuhan in Hubei, China, reported a series of cases of pneumonia with unknown aetiology ^1^. The Chinese Centre for Disease Control and Prevention (CCDC) described severe acute respiratory syndrome-related coronavirus-2 (SARS-CoV-2) as the causative agent ^2-4^, which then quickly spread worldwide. Severe cases of SARS-CoV-2 infection are associated with a substantial risk of prolonged critical illness and death^5^.

Because the virus can be spread by asymptomatic, pre-symptomatic, and symptomatic carriers, public health experts have recommended fast and accurate testing, followed by the identification and monitoring of positive cases and subsequent self-isolation and contact tracing to contain the spread of SARS-CoV-2. Direct detection of SARS-CoV-2 is achieved by identifying viral RNA in specimens from the respiratory tract of patients utilizing nucleic acid amplification tests (NAATs) or by recognising viral proteins through antigen-detecting rapid diagnostic tests (Ag-RDTs). The NAAT-based assays, such as real-time reverse transcription-polymerase chain reaction (rRT-PCR) have become the “gold standard” for establishing the presence of SARS-CoV-2 in patients ^6^. However, rRT-PCR-based testing takes several hours and is conducted in specialised laboratories, which are usually located away from sample collection sites. This may produce long turnaround times, resulting in delayed self-isolation, a risk of more contacts, and further potential transmission. Therefore, rapid antigen tests for SARS-CoV-2 detection have been made commercially available. Ag-RDTs can be conducted at the point of care and the results visualised after 15-30 minutes ^7^. There is common consensus that positive Ag-RDT results must be verified by rRT-PCR testing. Studies have indicated that the antigen tests’ analytical sensitivities vary between 25% and 50% for rRT-PCR-positive samples, which may increase to more than 80% for samples with a higher viral load ^8-10^. We set out to comprehensively examine the most sensitive ^11^ and widely used commercial RDTs in a real-world, prospective, head-to-head study, placing specific emphasis on clinical characteristics, COVID-19-associated symptoms, and the presence of SARS-CoV-2 variant genotypes.

## Methods

### Setting and participants

This prospective study was conducted at the Corona Test Centre Stuttgart Cannstatter Wasen. Individuals scheduled for rRT-PCR testing of nasopharyngeal swabs were advised of the study orally and in writing. Participants had to be aged ≥18 years and capable of understanding the nature, significance, and implications of the study. Children and adolescents <18 years of age and patients obviously suffering from clinical conditions requiring emergency hospitalisation were excluded. All participants provided written and informed consent. The study was approved by Ethics Committee II (Mannheim) of the University of Heidelberg (reference number 2020-417MF) and the German Institute for Drugs and Medical Devices.

We recorded demographic characteristics, reasons for testing, medical history, major risk factors, acute symptoms, and vital signs, including heart rate, blood pressure, body temperature, and oxygen saturation (Table 1). In addition to collecting the nasopharyngeal swabs for rRT-PCR testing, we collected two completely independent swab specimens to run two commercially available and widely used Ag-RDTs from Abbott Diagnostics (Abbott Panbio **TM** COVID-19 Ag Rapid Test, Abbott Rapid Diagnostics Jena GmbH, Jena, Germany, www.abbott.com/poct) and Roche Diagnostics (Roche-SD Biosensor SARS-CoV-2 Rapid Antigen Test, identical to SD BIOSENSOR Standard Q COVID-19 Ag, www.sdbiosensor.com; Roche Diagnostics; Mannheim, Germany, www.roche.com). Hereafter, we refer to the tests as Abbott-RDT and Roche-RDT, respectively. We randomly assigned the study population to three sampling groups according to the time sequence of collecting the nasopharyngeal swabs (group 1: rRT-PCR, RDT-Roche, RDT-Abbott; group 2: RDT-Roche, RDT-Abbott, rRT-PCR; and group 3: RDT-Abbott, rRT-PCR, RDT-Roche).

**Table 1.**
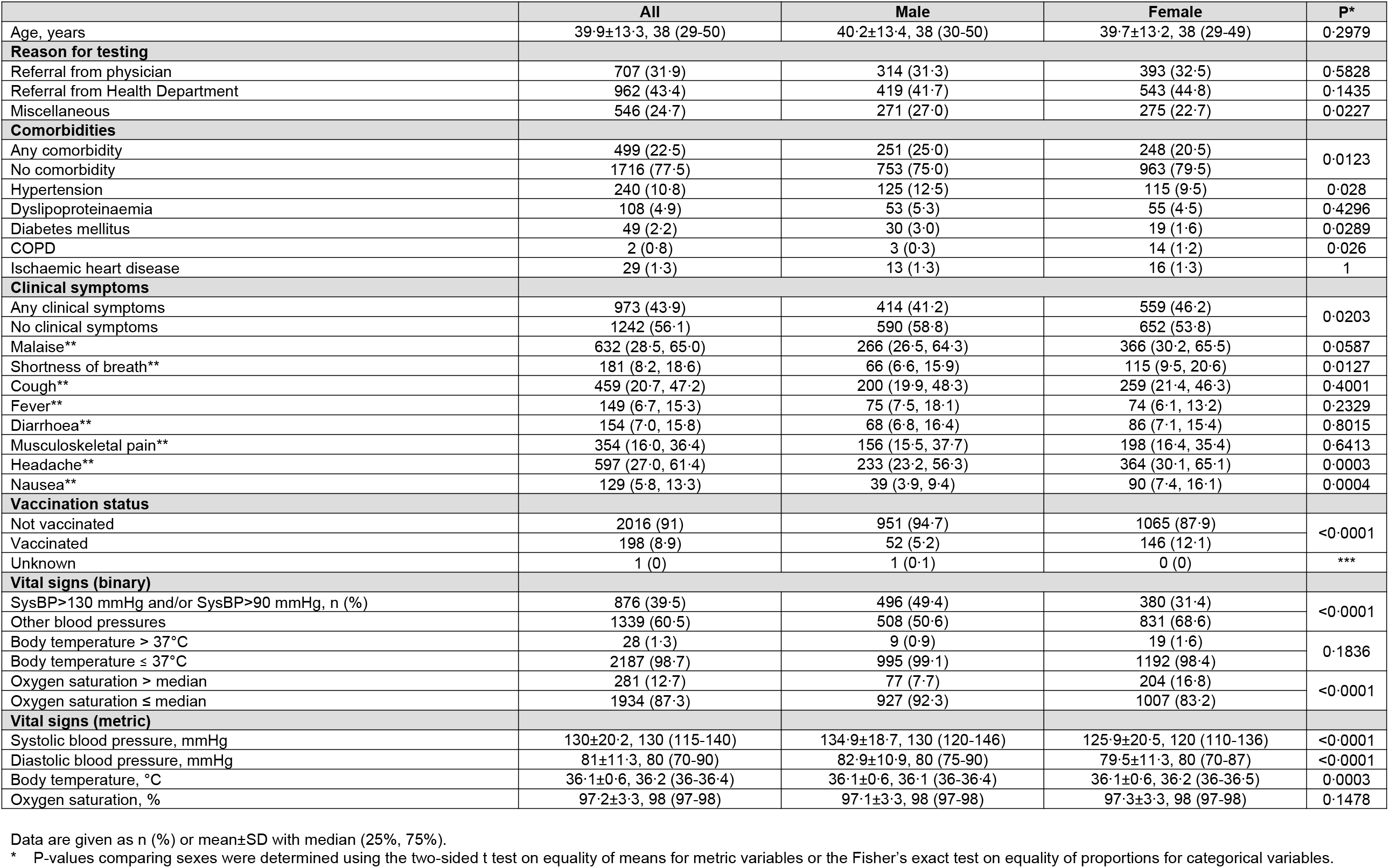
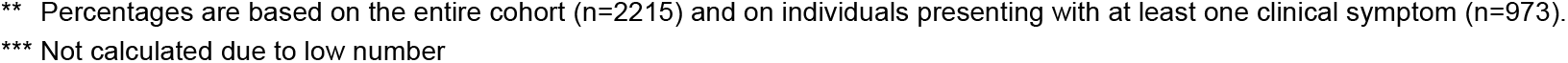
Clinical characteristics of the 2215 participants in the COVAG study.

|  | All | Male | Female | P* |
| --- | --- | --- | --- | --- |
| Age, years | 39.9±13.3, 38 (29-50) | 40.2±13.4, 38 (30-50) | 39.7±13.2, 38 (29-49) | 0.2979 |
| **Reason for testing** |  |  |  |  |
| Referral from physician | 707 (31.9) | 314 (31.3) | 393 (32.5) | 0.5828 |
| Referral from Health Department | 962 (43.4) | 419 (41.7) | 543 (44.8) | 0.1435 |
| Miscellaneous | 546 (24.7) | 271 (27.0) | 275 (22.7) | 0.0227 |
| **Comorbidities** |  |  |  |  |
| Any comorbidity | 499 (22.5) | 251 (25.0) | 248 (20.5) | 0.0123 |
| No comorbidity | 1716 (77.5) | 753 (75.0) | 963 (79.5) |  |
| Hypertension | 240 (10.8) | 125 (12.5) | 115 (9.5) | 0.028 |
| Dyslipoproteinaemia | 108 (4.9) | 53 (5.3) | 55 (4.5) | 0.4296 |
| Diabetes mellitus | 49 (2.2) | 30 (3.0) | 19 (1.6) | 0.0289 |
| COPD | 2 (0.8) | 3 (0.3) | 14 (1.2) | 0.026 |
| Ischaemic heart disease | 29 (1.3) | 13 (1.3) | 16 (1.3) | 1 |
| **Clinical symptoms** |  |  |  |  |
| Any clinical symptoms | 973 (43.9) | 414 (41.2) | 559 (46.2) | 0.0203 |
| No clinical symptoms | 1242 (56.1) | 590 (58.8) | 652 (53.8) |  |
| Malaise** | 632 (28.5, 65.0) | 266 (26.5, 64.3) | 366 (30.2, 65.5) | 0.0587 |
| Shortness of breath** | 181 (8.2, 18.6) | 66 (6.6, 15.9) | 115 (9.5, 20.6) | 0.0127 |
| Cough** | 459 (20.7, 47.2) | 200 (19.9, 48.3) | 259 (21.4, 46.3) | 0.4001 |
| Fever** | 149 (6.7, 15.3) | 75 (7.5, 18.1) | 74 (6.1, 13.2) | 0.2329 |
| Diarrhoea** | 154 (7.0, 15.8) | 68 (6.8, 16.4) | 86 (7.1, 15.4) | 0.8015 |
| Musculoskeletal pain** | 354 (16.0, 36.4) | 156 (15.5, 37.7) | 198 (16.4, 35.4) | 0.6413 |
| Headache** | 597 (27.0, 61.4) | 233 (23.2, 56.3) | 364 (30.1, 65.1) | 0.0003 |
| Nausea** | 129 (5.8, 13.3) | 39 (3.9, 9.4) | 90 (7.4, 16.1) | 0.0004 |
| **Vaccination status** |  |  |  |  |
| Not vaccinated | 2016 (91) | 951 (94.7) | 1065 (87.9) | <0.0001 |
| Vaccinated | 198 (8.9) | 52 (5.2) | 146 (12.1) |  |
| Unknown | 1 (0) | 1 (0.1) | 0 (0) | *** |
| **Vital signs (binary)** |  |  |  |  |
| SysBP>130 mmHg and/or SysBP>90 mmHg, n (%) | 876 (39.5) | 496 (49.4) | 380 (31.4) | <0.0001 |
| Other blood pressures | 1339 (60.5) | 508 (50.6) | 831 (68.6) |  |
| Body temperature > 37°C | 28 (1.3) | 9 (0.9) | 19 (1.6) | 0.1836 |
| Body temperature ≤ 37°C | 2187 (98.7) | 995 (99.1) | 1192 (98.4) |  |
| Oxygen saturation > median | 281 (12.7) | 77 (7.7) | 204 (16.8) | <0.0001 |
| Oxygen saturation ≤ median | 1934 (87.3) | 927 (92.3) | 1007 (83.2) |  |
| **Vital signs (metric)** |  |  |  |  |
| Systolic blood pressure, mmHg | 130±20.2, 130 (115-140) | 134.9±18.7, 130 (120-146) | 125.9±20.5, 120 (110-136) | <0.0001 |
| Diastolic blood pressure, mmHg | 81±11.3, 80 (70-90) | 82.9±10.9, 80 (75-90) | 79.5±11.3, 80 (70-87) | <0.0001 |
| Body temperature, °C | 36.1±0.6, 36.2 (36-36.4) | 36.1±0.6, 36.1 (36-36.4) | 36.1±0.6, 36.2 (36-36.5) | 0.0003 |
| Oxygen saturation, % | 97.2±3.3, 98 (97-98) | 97.1±3.3, 98 (97-98) | 97.3±3.3, 98 (97-98) | 0.1478 |

Data are given as n (%) or mean±SD with median (25%, 75%).
\* P-values comparing sexes were determined using the two-sided t test on equality of means for metric variables or the Fisher's exact test on equality of proportions for categorical variables.
\*\* Percentages are based on the entire cohort (n=2215) and on individuals presenting with at least one clinical symptom (n=973).
\*\*\* Not calculated due to low number

Blood samples (two 7·5 ml S-Monovette^®^ Serum-Gel tubes and a 2·7 ml S-Monovette^®^ K_3_-EDTA tube, Sarstedt, Nümbrecht, Germany) were collected from the participants in a resting position to create a biobank. These backup samples were stored at -80 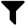 C for future experiments. The full study protocol is available through https://www.synlab.de/human/news-artikel/veroeffentlichung-des-protokolls-der-covid-19-antigen-studie-1398.

### Analytical procedures

Both the Abbott-RDT and the Roche-RDT were carried out by medically educated staff according to the manufacturers’ instructions on-site at the Corona Test Centre Cannstatter Wasen, Stuttgart, Germany, immediately after sampling the nasopharyngeal swabs. The nasopharyngeal swabs for rRT-PCR testing were placed in 2 ml of a phosphate-buffered saline solution (ISOTON™ II Diluent, Becton Dickinson, Galway, Ireland) and delivered to the SYNLAB Medical Care Centre Leinfelden-Echterdingen. This ensured that the performers of the RDTs were unaware of the rRT-PCR-results.

SARS-CoV-2 RNA was extracted from the nasopharyngeal swab samples and purified using the PurePrep Pathogens kit and a PurePrep 96 instrument (Molgen, Veenendaal, the Netherlands). The rRT-PCR assay was performed using the RIDA^®^GENE SARS-CoV-2 test kit (R-Biopharm, Darmstadt, Germany) on the CFX96 Touch Real-Time PCR detection device (Bio-Rad, Feldkirchen, Germany) according to the manufacturers’ instructions. This test kit targets the SARS-CoV-2 2 envelope (E) gene; samples producing a cycle threshold (Ct) ≤ 35 were considered positive by rRT-PCR.

We screened rRT-PCR-positive samples for SARS-CoV-2 variants of concern (VOC) B.1.1.7 (Alpha, United Kingdom), B.1.351 (Beta, South Africa), and P.1 (Gamma, Brazil) using VirSNiP SARS-CoV-2 Spike N501Y and VirSNiP SARS-CoV-2 Spike del H69/V70 from TIB Molbiol (Berlin, Germany) according to the supplier’s instructions. Genotyping was restricted to samples with a Ct ≤ 30 in rRT-PCR (260 of 338 samples). Samples with positive results for both the N501Y substitution and H69/V70 deletion were assigned to the Alpha variant. The presence of N501Y and absence of H69/V70 was considered the Beta or Gamma variant. Samples in which both N501Y and H69/V70 were absent may have contained variants other than Alpha, Beta, or Gamma or the wild-type. For the purpose of this article, these samples were termed negative for investigated mutations (NIM).

### Statistical analysis

Continuous data are presented as the mean, standard deviation (SD), median, and 25^th^ and 75^th^ percentiles. Categorical data are presented as absolute numbers and percentages. Differences in continuous variables between men and women were examined by two-sided t-tests; the chi-squared test was used for categorical variables (Table 1). The risk of having COVID-19 according to baseline anthropometric and anamnestic characteristics was expressed in terms of crude odds ratios (ORs) and ORs adjusted for age and sex as calculated by logistic regression (Table 2). Sensitivity, specificity, positive predictive values (PPVs), negative predictive values (NPVs), and diagnostic efficacy of the two RDTs compared to rRT-PCR were calculated (Table 3). These performance indicators were compared between the Abbott-RDT and Roche-RDT. The P-value refers to two-sided testing of the null hypothesis, that the difference between the respective performance indicators is equal to zero and based on 5000 bootstrap iterations and subsequent percentile analysis. We also visualised the sensitivities of both RDTs relative to the rRT-PCR-derived Ct values (Figure 1) and the PPVs and NPVs according to hypothetical disease prevalence rates in the range of 0-0·05 (Figure 2). Finally, we investigated whether the sensitivities of the two RDTs were related to the reason for testing, comorbidities, clinical symptoms, vital signs, or SARS-CoV-2 genotypes using univariate (Table 3) and multivariate logistic regression (Table 4).

**Table 2.** Risk for rRT-PCR-proven SARS2-Cov-2 infection according to clinical characteristics in 2215 participants of the COVAG study.

|  | N | PCR Positive, N (%) | OR (95% CI) | OR (95% CI) |
| --- | --- | --- | --- | --- |
|  |  |  | Crude | Adjusted for age and sex |
| Age (per year) * |  |  | 1 (0.99, 1.01) |  |
| Male sex (reference: female) | 1004 | 164 (16.3) | 1.16 (0.92, 1.47) |  |
| **REASON FOR TESTING** |  |  |  |  |
| Referral from physician | 707 | 122 (17.3) | 1.0 (reference) | 1.0 (reference) |
| Referral from Health Department (mostly contact persons of infected patients) | 962 | 192 (20) | 1.2 (0.93, 1.54) | 1.2 (0.93, 1.54) |
| other | 546 | 24 (4.4) | 0.22 (0.14, 0.35) | 0.22 (0.14, 0.34) |
| **COMORBIDITIES** |  |  |  |  |
| Any comorbidity | 499 | 131 (26.3) | 2.6 (2.03, 3.32) | 2.94 (2.25, 3.83) |
| No comorbidity | 1716 | 207 (12.1) | 1.0 (reference) | 1.0 (reference) |
| Hypertension | 240 | 37 (15.4) | 1.33 (0.91, 1.94) | 1.46 (0.97, 2.2) |
| Dyslipoproteinaemia | 108 | 18 (16.7) | 1.46 (0.86, 2.47) | 1.76 (1.01, 3.05) |
| Diabetes mellitus | 49 | 10 (20.4) | 1.87 (0.92, 3.8) | 2.24 (1.08, 4.63) |
| COPD | 17 | 2 (11.8) | 0.97 (0.22, 4.28) | 1.19 (0.27, 5.27) |
| Ischaemic heart disease | 29 | 7 (24.1) | 2.32 (0.98, 5.5) | 2.87 (1.19, 6.91) |
| Previous COVID-19 | 182 | 99 (54.4) | 8.7 (6.28, 12.04) | 8.61 (6.21, 11.93) |
| **CLINICAL SYMPTOMS** |  |  |  |  |
| Any clinical symptoms | 973 | 222 (22.8) | 2.87 (2.25, 3.66) | 2.91 (2.28, 3.71) |
| No clinical symptoms | 1242 | 116 (9.3) | 1.0 (reference) | 1.0 (reference) |
| Malaise | 632 | 155 (24.5) | 3.15 (2.42, 4.1) | 3.2 (2.46, 4.17) |
| Shortness of breath | 181 | 48 (26.5) | 3.5 (2.39, 5.13) | 3.6 (2.45, 5.28) |
| Cough | 459 | 133 (29) | 3.96 (3, 5.23) | 4.01 (3.03, 5.3) |
| Fever | 149 | 58 (38.9) | 6.19 (4.23, 9.05) | 6.17 (4.22, 9.04) |
| Diarrhoea | 154 | 32 (20.8) | 2.55 (1.65, 3.93) | 2.58 (1.67, 3.98) |
| Musculoskeletal pain | 354 | 114 (32.2) | 4.61 (3.44, 6.18) | 4.67 (3.48, 6.26) |
| Headache | 597 | 128 (21.4) | 2.65 (2.02, 3.48) | 2.7 (2.05, 3.56) |
| Nausea | 129 | 25 (19.4) | 2.33 (1.45, 3.76) | 2.42 (1.5, 3.91) |
| **VACCINATION STATUS** |  |  |  |  |
| Not vaccinated | 2016 | 323 (16) | 2.33 (1.36, 3.99) | 2.28 (1.32, 3.92) |
| Vaccinated | 198 | 15 (7.6) | 1.0 (reference) | 1.0 (reference) |
| Unknown | 1 | 0 (0) | ** | ** |
| **VITAL SIGNS** |  |  |  |  |
| SysBP>130 mmHg and/or SysBP>90 mmHg | 876 | 125 (14.3) | 0.88 (0.69, 1.12) | 0.84 (0.65, 1.09) |
| Body temperature > 37°C | 28 | 12 (42.9) | 4.28 (2.01, 9.13) | 4.4 (2.05, 9.41) |
| Oxygen saturation > median | 281 | 38 (13.5) | 0.85 (0.59, 1.22) | 0.87 (0.6, 1.26) |

\* For metric variables, the estimated increase in log-odds for rRT-PCR-proven SARS2-Cov-2 infection per one-unit increase.
\*\* Not calculated due to low number

**Table 3.**
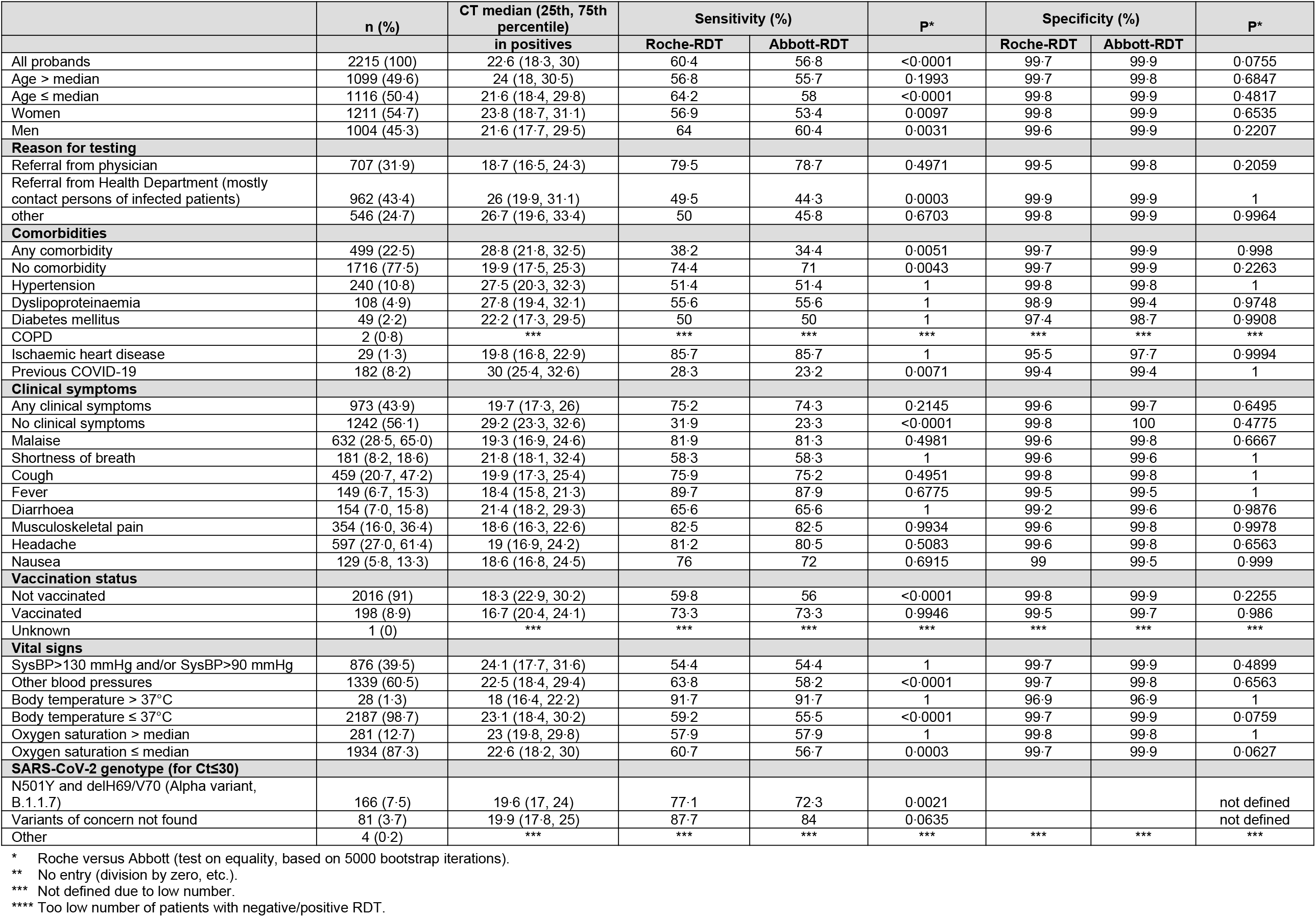
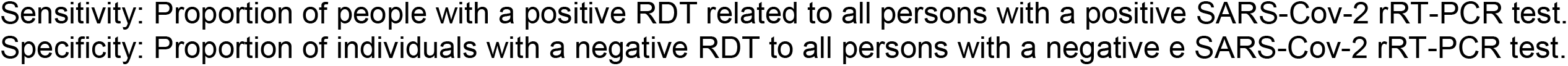
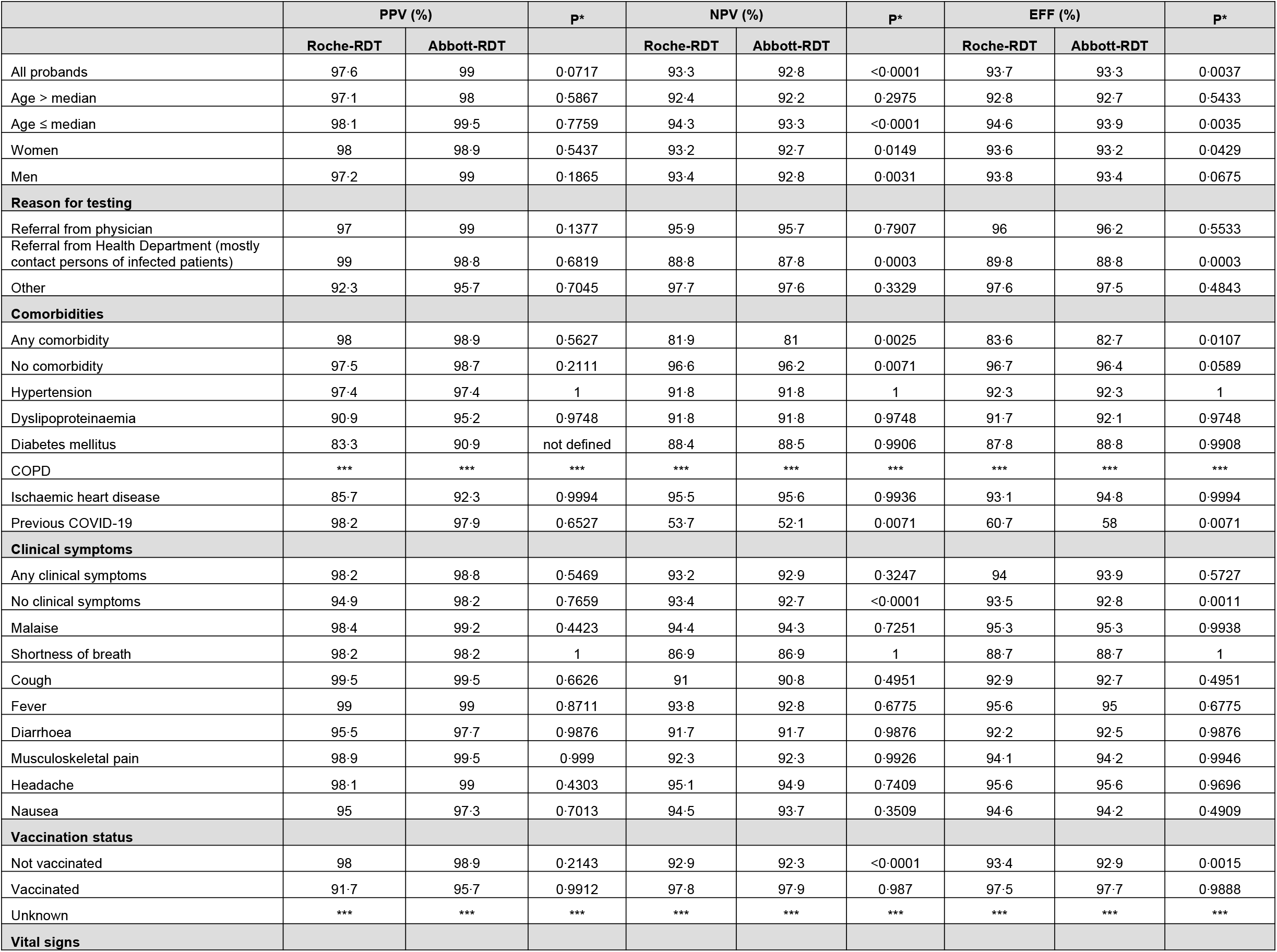
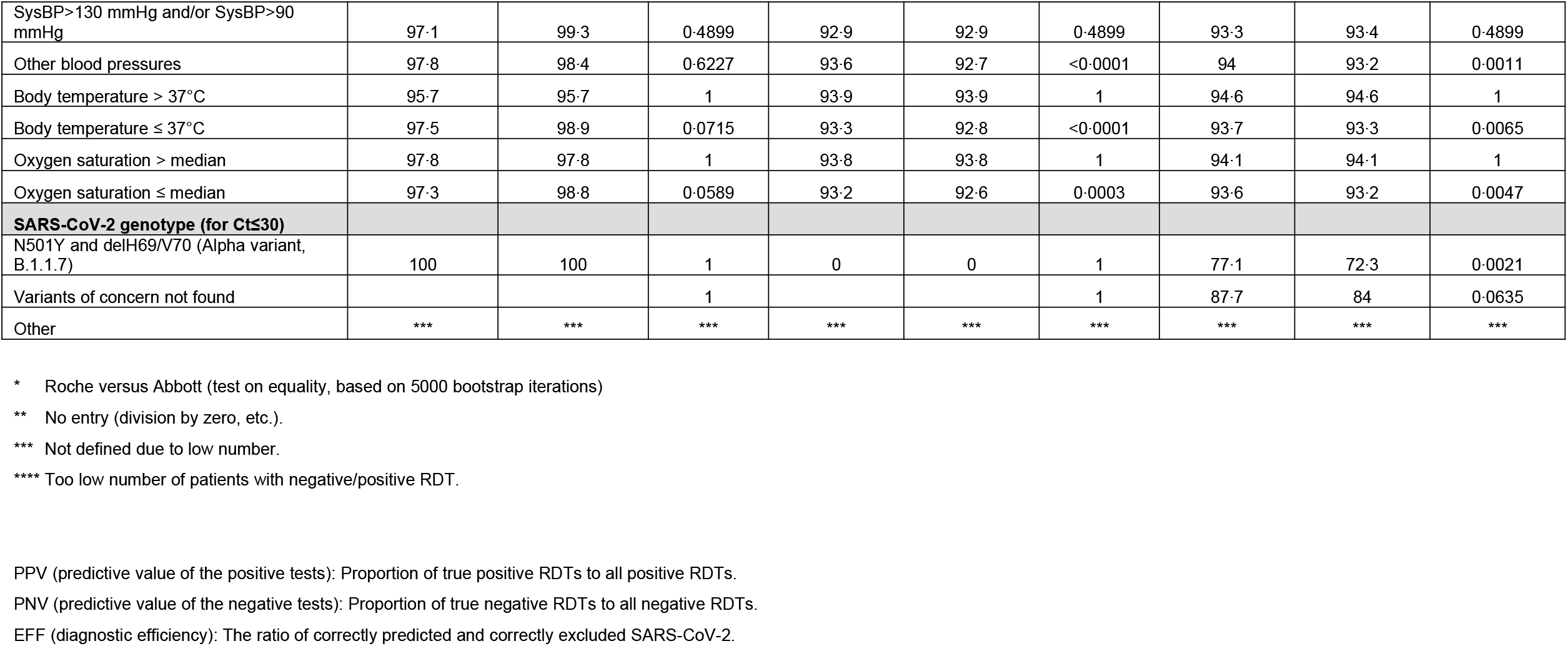
Diagnostic performance of two commercial RDTs for SARS-Cov-2 antigen.

**Figure 1.**
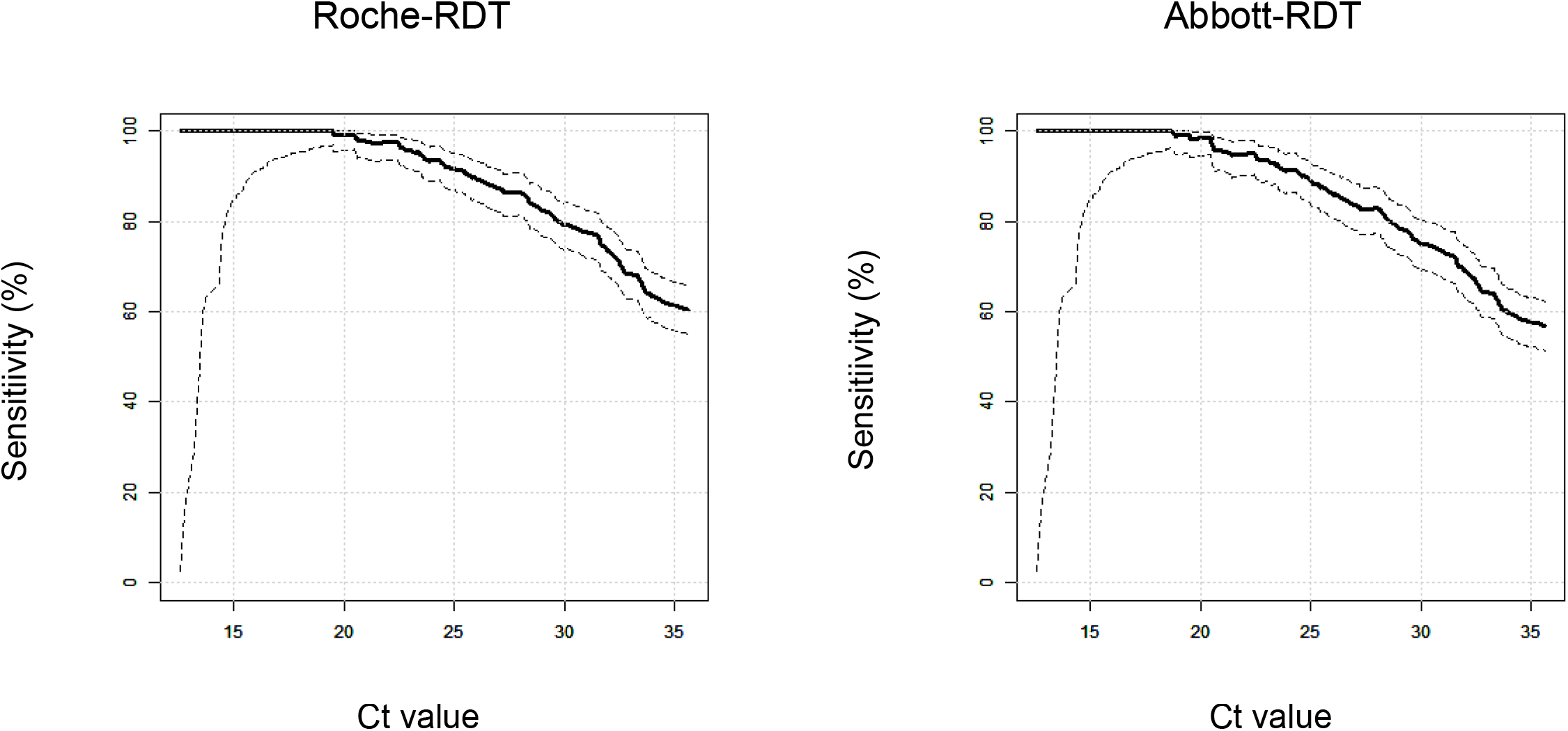
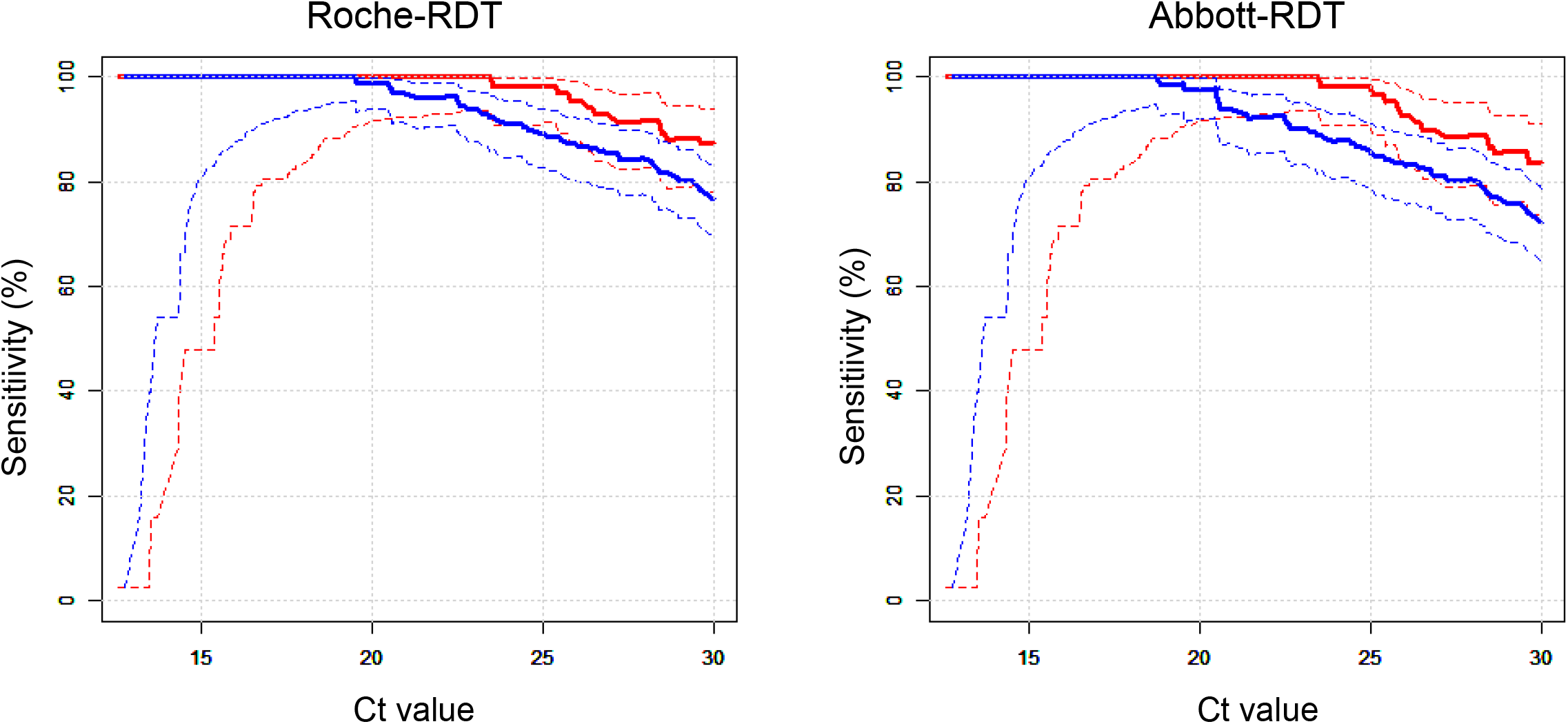
Cycle threshold (Ct) values in rRT-PCR for SARS-Cov-2 RNA versus the sensitivities of the Roche-RDT (*left*) and Abbott-RDT (*right*). *A*. The solid lines indicate sensitivities, the dotted lines represent the upper and lower bounds of the corresponding 95% confidence intervals. *B*. The solid lines indicate sensitivities, the dotted lines represent the upper and lower bounds of the corresponding 95% confidence intervals. Red: SARS-CoV-2 NIM genotype; blue: SARS-CoV-2 Alpha variant.

**Figure 2.**
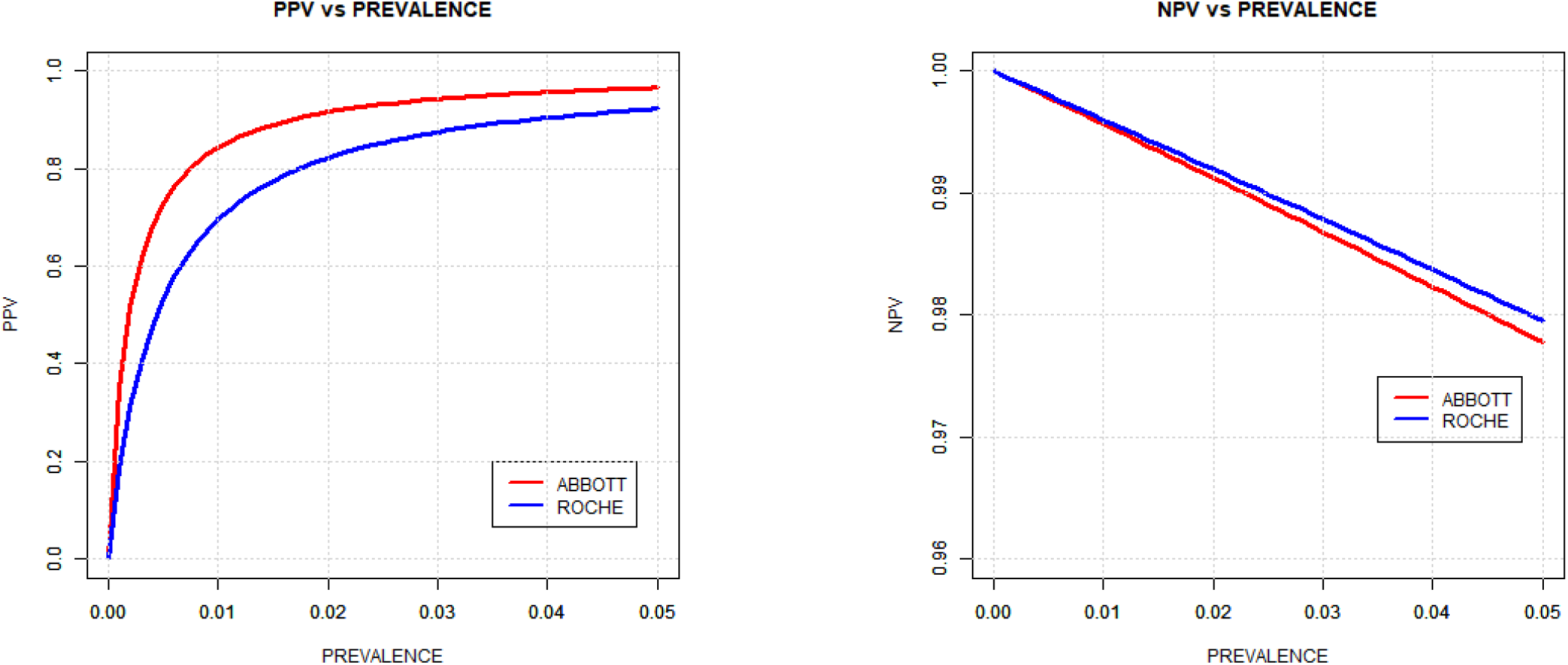
Positive predictive values (PPVs, right *panel*) and negative predictive values (NPVs, *left panel*) of two commercial RDTs for SARS-CoV-2-associated antigens in relation to disease prevalence rates up to 0·05. *Red line:* Abbott-RDT; *blue line:* Roche-RDT.

**Table 4.** Predictors of positive RDTs amongst SARS-CoV-2-positive samples in a multivariate model.

|  |  | Roche-RDT |  | Abbott-RDT |  |
| --- | --- | --- | --- | --- | --- |
| Covariable |  | OR (95% CI) | P | OR (95% CI) | P |
| Age and sex | Age, per year | 0·99 (0·96, 1·02) | 0·557 | 1·03 (0·99, 1·07) | 0·098 |
|  | Men | 1·0 (reference) |  | 1·0 (reference) |  |
|  | Women | 1·14 (0·46, 2·81) | 0·776 | 0·79 (0·32, 1·94) | 0·605 |
| Ct value | Ct value on rRT-PCR, per unit | 0·68 (0·6, 0·78) | <0·0001 | 0·64 (0·56, 0·74) | <0·0001 |
| Reason for testing | Referral from physician | 1·0 (reference) |  | 1·0 (reference) |  |
|  | Referral from Health Department | 0·53 (0·16, 1·71) | 0·287 | 0·54 (0·17, 1·72) | 0·297 |
|  | Miscellaneous reasons | 1·95 (0·1, 39·67) | 0·665 | 0·87 (0·07, 10·66) | 0·914 |
| Comorbidities | No comorbidity | 1·0 (reference) |  | 1·0 (reference) |  |
|  | Any comorbidity | 0·89 (0·34, 2·33) | 0·807 | 0·81 (0·29, 2·24) | 0·681 |
| Clinical symptoms | No clinical symptoms | 1·0 (reference) |  | 1·0 (reference) |  |
|  | At least one clinical symptom | 2·22 (0·91, 5·38) | 0·079 | 5·51 (2·18, 13·91) | 0·0003 |
| SARS-CoV-2 genotype | SARS-CoV-2 NIM | 1·0 (reference) |  | 1·0 (reference) |  |

The statistical tests were two-sided and P<0·05 was considered significant. The analyses were carried out using R v4.0.2 (http://www.r-project.org).

## Results

### Clinical characteristics of participants

The study was conducted between February 1, 2021, and March 31, 2021. During this period, nearly 17 000 adult persons attended the test centre Cannstatter Wasen to receive an rRT-PCR test. A total of 2222 persons agreed to participate in the study. Seven of them were disregarded for further evaluation because at least one of the three tests was not available. This resulted in 2215 persons with valid data (Table 1). Adverse events from performing any of the tests were not experienced.

We divided the reasons for testing into three major categories (Table 1): 707 (32%) participants were referred by their primary care physicians, 962 (43%) sought testing following the advice of the Health Department and were mostly incriminated contact persons of COVID-19 patients, and 546 (25%) participants were tested for miscellaneous reasons (264 following a warning message from the German Corona App, 140 out-of-pocket payers, 82 kindergartners or teachers, and 60 for another reason, including 2 for confirmation of a positive RDT and 2 cluster students in quarantine).

Hypertension, dyslipoproteinaemia, and diabetes mellitus were self-reported at rates of 12%, 5%, and 2%, respectively. Chronic obstructive lung disease and ischaemic heart disease were comparatively low in frequency. Overall, comorbidities occurred more often in men than in women (Table 1).

The most often reported clinical symptoms were malaise, headache, and musculoskeletal pain (Table 1). Symptoms were significantly more frequent in women than in men. Systolic and diastolic blood pressures were markedly and significantly higher in men than in women, but body temperature was slightly higher in women than in men (Table 1).

### Risk of rRT-PCR-proven SARS-CoV-2 infection according to baseline characteristics

Among the 2215 participants, 338 carried SARS-CoV-2 based on rRT-PCR. Age and sex were not related to the likelihood of testing positive for SARS-CoV-2 (Table 2). Participants referred by treating physicians and health departments were positive significantly more often than participants with miscellaneous reasons for testing (OR 0·22, 95% confidence interval [CI] 0·14-0·34, adjusted for age and sex and compared to referrals from physicians).

Persons with at least one comorbidity more frequently tested positive than those without comorbidity (OR 2·94, 95% CI 2·25-3·83, adjusted for age and sex). Among the individual comorbidities, dyslipoproteinaemia, diabetes mellitus, and ischaemic heart disease significantly increased the probability of a positive rRT-PCR test. The presence of at least one clinical symptom at presentation resulted in a higher frequency of positive rRT-PCR (OR 2·91, 95% CI 2·28-3·71, adjusted for age and sex). In addition, each of the individual symptoms was positively and significantly related to the rate of SARS-CoV-2 detected by rRT-PCR. Among the objectively measured vital signs (blood pressure, body temperature, and oxygen saturation), elevated body temperature was associated with the probability of COVID-19.

### Diagnostic performance of RDTs

#### Sensitivity

The Roche-RDT and Abbott-RDT had overall sensitivities of 60·4% and 56·8%, respectively (P<0·0001, Table 3). Figure 1A shows that the sensitivities of both RDTs were strongly related to the Ct values derived from rRT-PCR. Only at Ct values < 20 did both RDTs reach a sensitivity of 100%.

We further examined whether the sensitivities of the two RDTs were significantly different in subgroups (Table 3). Age did not significantly affect sensitivity, but the overall difference in sensitivity between the Roche-RDT and Abbott-RDT may be due to participants below the median age (P<0·0001) rather than those above the median age (P=0·199, Table 3).

Among participants referred by physicians, the sensitivities of both RDTs were substantially higher than in the total study population (79·5% and 78·7%, Roche-RDT and Abbott-RDT, respectively) but did not differ significantly. In contrast, they were <50% in persons referred by the Health Department and those tested for other reasons, whereby the Roche-RDT appeared to perform better than the Abbott-RDT (Table 3).

Sensitivities were markedly lower in persons with at least one comorbidity (38·2% and 34·4%, Roche-RDT and Abbott-RDT, respectively, P=0·005) than in persons without comorbidities (74·4% and 71·0%, Roche-RDT and Abbott-RDT, respectively, P=0·004, Table 3). This also applied to the individual comorbidities, with the one exception that the sensitivities of both the Roche-RDT and Abbott-RDT were 86% in the small number of participants reporting ischaemic heart disease. This unexpected finding is consistent with the Ct values being markedly higher in individuals with comorbidities (Table 3).

In persons with at least one clinical symptom, the sensitivities of both RDTs were higher (75·2% and 74·3%, Roche-RDT and Abbott-RDT, respectively, not significant) than in persons without clinical symptoms (31·9% and 23·8%, Abbott-RDT and Roche-RDT, respectively, P<0·0001). The presence of any of single symptom augmented the sensitivities of both RDTs, with the exception of shortness of breath and diarrhoea, which were not related to sensitivity. This finding is in line with the Ct values, which were lower in cases with symptoms than in those without (Table 3). In the small number of persons (n = 12) with elevated body temperature, the rate of positive RDTs was higher (91·7% for both RDTs) than at normal body temperature (59·2% and 55·5%, Abbott-RDT and Roche-RDT, respectively, P<0·001), but there were only modest differences in sensitivities due to blood pressure and oxygen saturation (Table 3).

We also analysed whether the SARS-CoV-2 genotype affects the sensitivities of the RDTs. Only samples with Ct values ≤ 30 (n = 286) were included in this evaluation. The NIM SARS-CoV-2 and Alpha variant (B.1.1.7) were present in 81 and 166 samples with Ct values ≤ 30, respectively. The NIM genotype was detected at sensitivities of 87·7% and 84·0% (Roche-RDT versus Abbott-RDT, respectively, not significant). In carriers of the Alpha variant, sensitivities were 77·1% and 72·3% (Roche-RDT versus Abbott-RDT, respectively, P<0·002). At any given Ct value, the sensitivities of both RDTs were lower for the Alpha variant than for the NIM genotype (Figure 1B).

To firmly establish independent predictors of sensitivity, we calculated ORs for having a positive RDT according to subgroups in multivariate logistic regression. Covariables were age, gender, reason for testing, presence or absence of any comorbidity, presence of absence of any clinical symptom, and the SARS-CoV-2 genotype. As expected, Ct values were associated with sensitivity for both tests. The sensitivities of the Abbott-RDT and Roche-RDT were higher in symptomatic than in asymptomatic individuals. Remarkably, the sensitivities of both tests were significantly lower for the Alpha variant than for the NIM genotype (Table 4).

#### Specificity

The rate of false-positive RDTs was low. With both RDTs, specificity exceeded 99% overall and mostly in all participant strata (Table 3).

#### PPV, NPV, and diagnostic efficacy

At a prevalence rate of 15% in the study population, the PPVs of the two RDTs were approximately 98% and within the range of 90 to 100% in all subgroups examined. The NPVs of the RDTs were approximately 93%. Diagnostic efficacy also ranged between 90 and 100%.

Because patients with SARS-CoV-2 infections were enriched in our study population compared to the general population, we examined the PPVs and NPVs at assumed prevalence rates up to 0·05 (Figure 2). At this prevalence rate, our results suggest a PPV and NPV of 96·6% and 97·8% for Abbott-RDT and 92·3% and 98% for Roche-RDT. In *symptomatic* persons, the respective figures were 93·6% and 98·7% for Abbott-RDT and 90·8% and 98·7% for Roche-RDT (Figure 2B). In asymptomatic persons, the respective figures were 100% and 96·1% for Abbott-RDT and 90·4% and 96·5% for Roche-RDT (Figure 2).

## Discussion

We completed one of the largest prospective evaluations of RDTs for SARS-CoV-2-associated antigens in a real-world environment to date. We evaluated the two most sensitive ^11^ contemporary lateral-flow devices provided by Roche Diagnostics and Abbott Diagnostics. We found that the Abbott-RDT and Roche-RDT had significantly different sensitivities, but they were inversely and strongly related to the rRT-PCR-derived Ct values. In unadjusted examinations, the RDTs had higher sensitivity in individuals referred from treating physicians and health departments than in those tested for other reasons, in individuals without comorbidities compared to those with comorbidities, in individuals presenting with clinical symptoms and fever, and in carriers of the SARS-CoV-2 NIM genotype compared to carriers of the Alpha variant. The associations between the presence of clinical symptoms or the SARS-CoV-2 genotype and sensitivity were robust against adjustments for the Ct values.

### Prevalence of SARS-CoV-2 infections in the study cohort

Among the attendees of our corona test centre, the rate of individuals testing positive for SARS-CoV-2 infection by rRT-PCR was 15%, which markedly exceeds the prevalence rate in the German population during the study (approximately 0·3% estimated on the basis of 7-day incidence rates during the study period). The probability of testing positive by rRT-PCR was not related to age and sex. This is in contrast to the expected over-representation of older people and may reflect pre-selection for persons having an evident clinical indication for testing. The probability of testing positive by rRT-PCR was strongly linked to individual reasons for testing, to the presence of comorbidities, clinical complaints, and elevated body temperature. For example, the persons referred by physicians due to suspected COVID-19 or those coming from the Health Department were more often positive than the group with miscellaneous reasons (kindergartners/teachers, out-of-pocket payers). Yet, in the latter group (prevalence rate 0·044), the proportion of SARS-CoV-2 carriers was still 10-fold higher than in the general population (assumed prevalence 0·003).

### Sensitivity of the RDTs

Expectedly, both RDTs had higher sensitivities in subgroups with high viral loads (referral by physicians and health departments, clinical symptoms). However, both sensitivities and viral loads were low in patients presenting with comorbidities. This is unexpected and may reflect a referral bias in the sense that the indication for testing is more frequent and earlier in patients at high risk for severe COVID-19.

The relationship between the RDTs’ analytical sensitivity and viral load is in line with reports of sensitivities between 24·3% and 50% for RT-PCR-positive samples, which increased up to 81·8% and 100% for samples with high viral loads (>6 log_10_ RNA copies/ml) ^8-10^. In specimens from the upper respiratory tract, SARS-CoV-2 RNA peaks with the beginning of symptoms around day 4, decreases steadily during the first 10 days after illness onset, and can be detected up to 20 days after the onset of symptoms ^12-15^. However, viral loads are low during the earlier stage of infection and in the second week after the onset of symptoms ^16,17^. Thus, the time window for the detection of SARS-CoV-2 by RDTs appears to be narrower than with rRT-PCR and may be confined to the acute phase of infection. Consistently, we found a lower sensitivity of RDTs in asymptomatic individuals, suggesting limited usefulness of RDTs for screening such individuals, even if it is repeated on a regular basis.

We want to emphasise that the RDTs were carried out by medically educated personnel with strict adherence to the instructions issued by the manufacturers, perhaps explaining the low rate of false-positive results. Yet, RDTs have been widely recommended for self-testing or for testing by lay persons. Indeed, when Ag-RDT self-testing results were evaluated in a comparative study among *symptomatic* outpatients, self-testing (including self-read-out) yielded a sensitivity of 82·5% compared to professional nasopharyngeal sampling and testing ^18^. The same study noted variations in the sensitivity of Ag-RDT self-testing depending on the viral load of the sample. High viral loads ≥7·0 log10 SARS-CoV-2 RNA copies/ml led to a sensitivity of 96·6%, whereas low viral loads <7·0 log10 SARS-CoV-2 RNA copies/ml had decreased sensitivity (45·6% for Ag-RDT self-use and 54·5% for Ag-RDT professional use) ^18^. Thus, it appears that, at low viral loads, as encountered in *asymptomatic* persons, self-administered RDT testing may be even less effective in reality than it was in the current study.

### Sensitivity for the SARS-CoV-2 Alpha variant

During the conduct of our study, the SARS-CoV-2 Alpha variant became the prevailing genotype in Southern Germany. Consequently, the study included 83 carriers of SARS-CoV-2 NIM, 166 carriers of the Alpha variant, and 4 patients with other viral genotypes (251 of 338 samples with Ct values ≤ 30). Remarkably, the sensitivities of both RDTs were lower in Alpha variant carriers, and this finding was robust against adjustments for the viral load expressed in terms of Ct values (Table 4). The Alpha variant may be differentiated from the wild-type by two key mutations in the Spike protein: the N501Y substitution within the receptor-binding domain and the H69/V70 deletion. It may be approximately 80% more transmissible than the wild-type ^19^ due to conformational changes increasing the Spike protein’s affinity for the angiotensin-converting enzyme 2 (ACE2) receptor ^20-22^. SARS-CoV-2 variants may also confer decreased binding of therapeutic antibodies and protection by vaccination, whereby the Alpha variant may display the smallest variation in antigenicity compared to other circulating variants. This raises the attractive possibility that the lower reactivity of the current RDTs for the Alpha variant was related to structural alterations in the epitope(s) recognised by the detecting antibodies. However, the antibodies incorporated in both RDTs recognise the nucleocapsid protein (N-protein) ^23^. In a laboratory-based investigation (virus suspended in cell culture medium and saliva), the performance of the Roche-RDT and Abbott-RDT was not affected by variants. This is in contrast to the current results, which were collected in a real-world setting. A potential explanation for the discrepancies may be that both the Spike protein and the N-protein tightly interact, and that conformational changes in the Spike protein may affect the three-dimensional structure and accessibility for antibodies against the N-protein ^24^. Finally, we cannot rule out that the structural changes in other variants may be even greater than in the Alpha variant ^25^, so that their effect on sensitivity may be even stronger. Thus, any validation of RDTs for SARS-CoV-2 would also have to be extended to known and future variants rather than limited to the wild-type SARS-CoV-2.

### Implications for screening

A recent meta-analysis issued by the Cochrane Collaboration reviewed 48 studies including 58 commercial RDTs. It reported sensitivities between 34·1% (95% CI 29·7% - 38·8%) and 88·1% (95% CI 84·2% - 91·1%) for RDTs in *symptomatic* persons, whereas in *asymptomatic* persons, the sensitivities varied between 28·6%(8·4% to 58·1%) and 69·2% (38·6% to 90·9%) ^11^.

The Cochrane analysis identified three studies on the Abbott-RDT with 1094 *symptomatic* participants, including 252 SARS-CoV-2 cases, and one study with 474 *asymptomatic* persons and 47 cases. It also identified three studies with 1948 *symptomatic* participants and 336 cases and one study with 127 *asymptomatic* persons and 13 cases for the Roche-RDT (listed as Biosensor Standard Q). The number of *asymptomatic* persons in the current study at least exceeds the number in the study in the Cochrane meta-analysis ^11^.

For the Abbott-RDT, the Cochrane analysis reported sensitivities of 75·1% (57·3% to 87·1%) and 48·9% (35·1% to 62·9%), and specificities of 99·5% (98·7% to 99·8%) and 98·1% (96·3% to 99·1%) in *symptomatic* and *asymptomatic* persons, respectively. The sensitivity of the Roche-RDT was reported to be 88·1% (84·2% to 91·1%) and 69·2% (38·6% to 90·9%) in *symptomatic* and *asymptomatic* patients, respectively, with specificities similar to the Abbott-RDT. The current findings almost exactly coincide with the Cochrane analysis and significantly extends the available evidence.

Analogous with the viewpoint of the Cochrane analysis ^11^, at a prevalence rate of 0·05, sensitivities of 60·4% and 56·8% and specificities of 99·7% and 99·9% with the Roche-RDT and Abbott-RDT, respectively, in *symptomatic* patients with SARS-CoV-2 infections, our data would translate as follows.

If 10 000 patients and 500 (0·05) true positives were examined, 414 and 397 persons would have tested positive, of which 38 and 25 would have been false-positives and 124 and 128 persons with negative test results would be falsely negative for Roche-RDT and Abbott-RDT, respectively (Table 5). Assuming a prevalence rate of 50 (0·005) in 10 000 *asymptomatic* patients, the respective figures would be 33 and 16 persons testing positive, of which 17 and 4 would have been false-positives for Roche-RDT and Abbott-RDT, respectively, and 34 and 38 persons with a negative test would be falsely negative. This is crucially important, as RDTs have specifically been recommended for screening asymptomatic persons (Table 5). Furthermore, as shown in Table 5, these figures would be substantially affected if the SARS-CoV-2 Alpha variant is predominant.

**Table 5.**
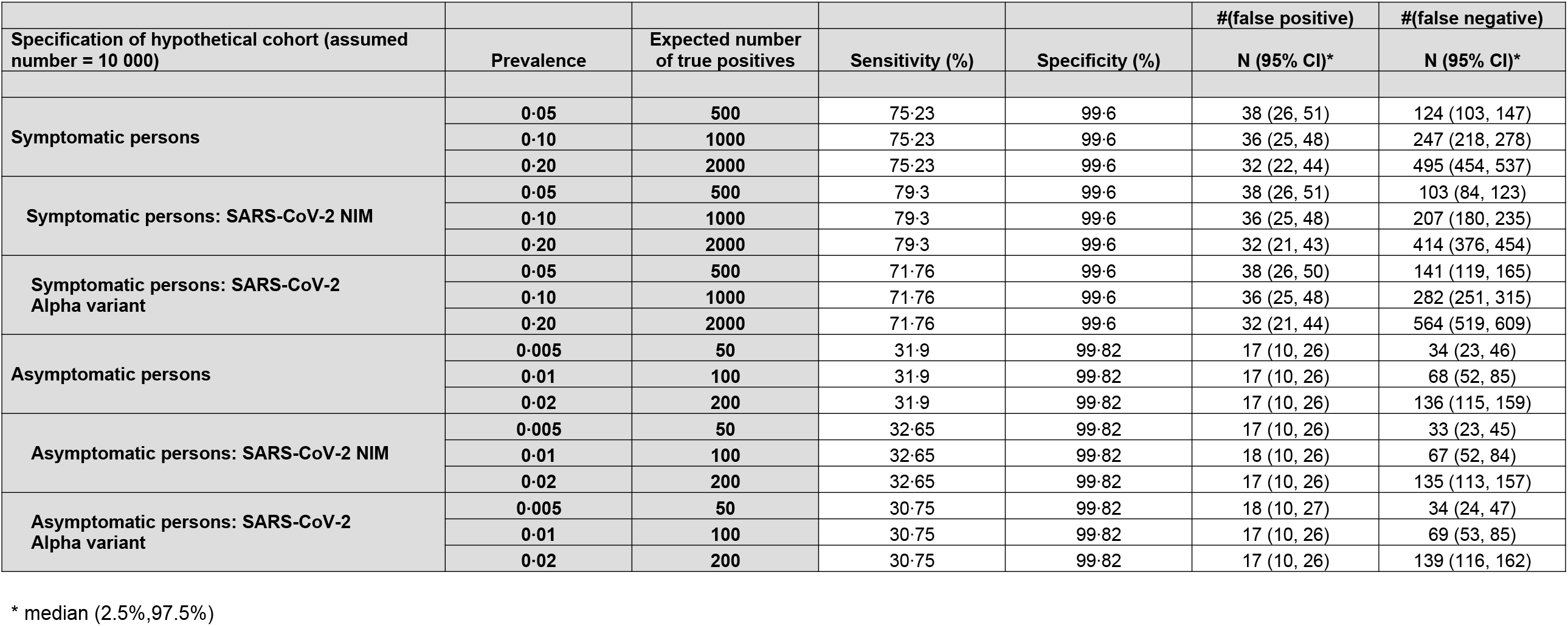
Number of false-positive and false-negative results in a hypothetical cohort of 10 000 people tested with the Roche-RDT.

**Table 5.** Number of false-positive and false-negative results in a hypothetical cohort of 10 000 people tested with the Abbott-RDT.

|  |  |  |  |  | #(false positive) | #(false negative) |
| --- | --- | --- | --- | --- | --- | --- |
| Specification of hypothetical cohort<br>(assumed number = 10 000) | Prevalence | Expected number<br>of true positives | Sensitivity (%) | Specificity (%) | N (95% CI)* | N (95% CI)* |
| Symptomatic persons | 0·05 | 500 | 74·32 | 99·73 | 25 (16, 36) | 128 (107, 151) |
|  | 0·10 | 1000 | 74·32 | 99·73 | 24 (15, 34) | 257 (227, 287) |
|  | 0·20 | 2000 | 74·32 | 99·73 | 21 (13, 31) | 513 (472, 556) |
| Symptomatic persons: SARS-CoV-2 NIM | 0·05 | 500 | 78·1 | 99·73 | 25 (16, 36) | 109 (90, 130) |
|  | 0·10 | 1000 | 78·1 | 99·73 | 24 (15, 34) | 219 (191, 248) |
|  | 0·20 | 2000 | 78·1 | 99·73 | 21 (13, 31) | 438 (398, 479) |
| Symptomatic persons: SARS-CoV-2<br>Alpha variant | 0·05 | 500 | 71·12 | 99·73 | 25 (16, 36) | 144 (121, 169) |
|  | 0·10 | 1000 | 71·12 | 99·73 | 24 (15, 34) | 289 (256, 322) |
|  | 0·20 | 2000 | 71·12 | 99·73 | 21 (13, 31) | 577 (532, 622) |
| Asymptomatic persons | 0·005 | 50 | 23·28 | 99·96 | 4 (1, 9) | 38 (27, 51) |
|  | 0·01 | 100 | 23·28 | 99·96 | 4 (1, 9) | 76 (60, 94) |
|  | 0·02 | 200 | 23·28 | 99·96 | 4 (1, 9) | 153 (130, 178) |
| Asymptomatic persons: SARS-CoV-2 NIM | 0·005 | 50 | 24·39 | 99·96 | 4 (1, 9) | 38 (27, 50) |
|  | 0·01 | 100 | 24·39 | 99·96 | 4 (1, 9) | 75 (59, 93) |
|  | 0·02 | 200 | 24·39 | 99·96 | 4 (1, 9) | 151 (128, 176) |
| Asymptomatic persons: SARS-CoV-2<br>Alpha variant | 0·005 | 50 | 21·58 | 99·96 | 4 (1, 9) | 39 (28, 52) |
|  | 0·01 | 100 | 21·58 | 99·96 | 4 (1, 9) | 78 (61, 96) |
|  | 0·02 | 200 | 21·58 | 99·96 | 4 (1, 9) | 157 (133, 183) |

\* median (2.5%,97.5%)

### Limitations

We applied rRT-PCR as the reference method to detect SARS-CoV-2 infection. Despite being considered the gold standard, this technique has the limitation that the detection of SARS-CoV-2 RNA in patient samples does not indicate the presence or shedding of viable virus with replicative capacity or whether the tested individual is contagious at the time of the test ^12,26^. Although the presence of viral RNA proven by rRT-PCR does not automatically equate to infectiousness, a significant correlation between the Ct value (reflecting viral load) and subsequent virus cultivation has been observed. Samples with Ct values between 13 and 17 have a culture positivity rate of 100%, which declines gradually to 12% when Ct values of 33 are analysed. No viral growth occurs at Ct values ≥34, suggesting that patients with these values do not excrete infectious viral particles ^17^. Therefore, if infectivity rather than a positive rRT-PCR test were considered the reference for RDTs, our results would stand more in favour of RDT testing. However, a direct conversion of Ct values or a positive RDT to contagiousness has not yet been established. Ct values can hardly be compared across studies, and the correlation between viral load and the risk of transmission from a positive case is still not entirely clear ^13,27^, with a variety of circumstances, such as the individual’s behaviour, the type and duration of contact, the environment, and the implementation of transmission-reducing measures (e.g., filter masks) affect infectiveness ^27,28^.

The sensitivity of RDTs may relate to the time elapsed since infection. It is a limitation of the current study that the time point of infection in rRT-PCR-positive samples was not available. However, we consider the Ct values from rRT-PCR as a good proxy for the changes in viral load during the course of the study.

Finally, we demonstrated that the SARS-CoV-2 Alpha variant markedly diminishes the sensitivity of both RDTs. We cannot explain this finding. Other variants were not encountered in sufficient numbers, and we were not able to infer the sensitivities of the RDTs for other variants.

### Directions for future research

This evaluation of the most sensitive RDTs currently available for SARS-CoV-2 suggests that screening asymptomatic persons with this approach may fail to identify a substantial proportion of viral carriers. Thus, further methodical refinements are needed, such as attempts to determine the viral load at least semi-quantitatively. Alternatively, rapid, on-site, direct detection of SARS-CoV-2 RNA RDTs could be pursued. The lower sensitivity of the RDTs for the Alpha variant indicates that their performance may be substantially influenced by the virus genotype, and strategies need to be developed to ensure that any of the circulating variants are captured. Finally, further head-to-head research is needed into how current screening strategies, RDT or laboratory-based, directly translate into controlling virus transmission and spread in the population. This will show whether the obvious practical advantage of RDTs is offset by their limitations.

## Data Availability

Data will be made available to researchers upon justified request and formal agreement to make sure that rules of good scientific practice are obeyed and that credit is given to the people who have been in charge of the design and the organization of the study. Interested researchers are invited to address their request or proposal to Prof. Dr. med. Winfried Maerz. The authors confirm that they accessed and validated these data and that all other researchers can access the data in the same manner the authors did.

## Data sharing

Data will be made available to researchers upon justified request and formal agreement to make sure that rules of good scientific practice are obeyed and that credit is given to the people who have been in charge of the design and the organization of the study. Interested researchers are invited to address their request or proposal to Prof. Dr. med. Winfried März. The authors confirm that they accessed and validated these data and that all other researchers can access the data in the same manner the authors did.

## Author contributions

WM, CHM, and CAM designed the study; CW, GBM, UH, and HJW collected the data; AD performed the statistical analysis; EW and GBM surveyed the laboratory analyses; CW and WM wrote the manuscript; all authors validated, reviewed, and edited the manuscript.

## Acknowledgements

We thank the participants for joining in free of remuneration. Katja Pöhl, CEO, SYNLAB MVZ Leinfelden-Echterdingen and Christoph Mahnke, CEO SYNLAB Holding Deutschland GmbH, supported the study. We thank Madeline Beckers, Alexander Ignatenko, Anna Süßmuth, Annika Saile, Berfin Cicek, Brigitte Schwandt, Carolin Meixner, Christian Lang, Felix Eisenmann, Jacques Pfander, Katerina Triantafillidi, Lars Banzhaf, Leo Rahmig, Miriam Berner, Robin Bohn, Susanne Kunz, Tim Fröschle, Verena Lebherz, and Jacob Göhring for carrying out the RDTs and documenting the participants at the Corona Test Centre Cannstatter Wasen, Stuttgart.

